# Reduced Grey Matter in Caudate and Accumbens Nuclei Precedes Central Post Stroke Pain

**DOI:** 10.64898/2025.12.08.25341786

**Authors:** Eleni Panagoulas, Karsten Mueller, Xiuhui Chen, Susanna Asseyer, Kersten Villringer, Thomas Krause, Gerhard Jan Jungehülsing, Arno Villringer

**Author notes:** Address correspondence to: Eleni Panagoulas and Arno Villringer.

## Abstract

Central Post Stroke Pain (CPSP) is a common, debilitating sequela of stroke that can occur within weeks to months after stroke onset, yet mechanistic insight and predictive biomarkers are lacking. To address this gap, we longitudinally studied patients with acute somatosensory stroke using structural and functional magnetic resonance imaging and assessed them for subsequent pain development. In the acute stage, patients who later developed CPSP had reduced grey matter concentration (GMC) in bilateral caudate nuclei and nuclei accumbens (NAc) and reduced functional connectivity from the contralesional caudate to the supplementary motor area (SMA) compared to those who remained pain-free. Reduced GMC of the contralesional caudate and reduced functional connectivity to SMA persisted after pain onset, furthermore, NAc exhibited reduced functional connectivity to bilateral sensorimotor cortices and SMA. Our findings provide evidence for neural changes that may predispose stroke patients to develop CPSP and point towards potential targets for pain prevention.

## Main text

Central post-stroke pain (CPSP) is a debilitating condition occurring in up to 50% of patients after a stroke affecting the somatosensory pathways [1]. As the incidence of stroke continues to rise, more patients are at risk of experiencing long-term sequelae such as CPSP. CPSP significantly reduces quality of life, affects patients’ rehabilitation progress and, given the large number of patients affected, places a significant burden on society in terms of increased demand on healthcare services [2].

The underlying mechanisms of pain development in CPSP are not well understood and treatment is often unsatisfactory, highlighting the urgency for early prediction and preventive intervention. To date, however, no biomarkers are available to predict which patients will develop CPSP after stroke [3]. Challenges in finding biomarkers and in developing effective treatment strategies include the high inter-individual variability seen in patients with CPSP [4] including the lesion itself, which can be located in the brainstem, thalamus, or somatosensory cortices. It is well established that thermal sensory dysfunction and damage to the spinothalamic tract (STT) or its projections are necessary but not sufficient for the development of CPSP. Central sensitisation is often suggested as the underlying mechanism of various forms of neuropathic pain [5]. However, it is unclear why only some stroke patients develop central sensitisation and experience chronic pain, while others with similar lesions and clinical profiles never develop pain [6].

One approach to answering this question has been to compare CPSP patients with stroke patients without pain, i.e., non-pain sensory stroke (NPSS), who are otherwise as similar as possible. While differences in lesion locations are very subtle in such comparative studies, employing computational neuroanatomy, e.g., voxel-based morphometry (VBM), it was shown that chronic CPSP patients have reduced grey matter in key pain-related brain areas (e.g. somatosensory, insular, and prefrontal cortices) compared to healthy controls [7]. Compared to NPSS controls, patients with CPSP showed reduced grey matter in contralesional S2, contralesional superior temporal as well as middle temporal gyrus and ipsilesional ventrolateral prefrontal cortex [7]. However, studies performed after pain has become chronic cannot distinguish between the precipitating factors that lead to the development of pain and the brain morphometric changes that accompany the pain. What is needed to possibly disentangle these factors are prospective longitudinal studies investigating participants before the onset of pain.

To address this we conducted a prospective longitudinal study. Patients were enrolled during the acute phase after stroke affecting the somatosensory system and were followed up for at least six months and up to one year. They were then classified as CPSP or NPSS according to standardised diagnostic criteria. We have recently reported the clinical data and results of detailed quantitative sensory testing in this sample [8]. To potentially identify neurobiological biomarkers that could predict the development of CPSP, 3 Tesla magnetic resonance imaging (MRI) was performed at baseline (“before pain”) and at follow-up (“with pain”). Both VBM based on structural MRI, and assessments of functional connectivity were performed to confirm VBM results with an independent approach and to provide additional information on the impact of structural neural changes on the functional connectome^1^.

Patients with acute stroke affecting the somatosensory system were prospectively enrolled at the Charité Universitätsmedizin Berlin, Germany (for details see also our previous publication [8]). After exclusion (decline or loss to follow-up, inadequate MRI quality or development of pain prior to the first MRI (see Supplementary Figure S1)), 65 patients (Table 1) were included in the imaging analysis “before pain”, comparing patients who subsequently developed pain (CPSP) and those who did not (NPSS). The groups did not differ significantly in terms of (sex CPSP F = 11, NPSS F = 13, *p* = 0.055) or age (CPSP median (min-max) = 65 (33-78) years, NPSS median (min-max) = 64 (24-80) years, *p* = 0.79 (W = 431) Table 1). The mean time from stroke to first MRI was 4.4 days and did not significantly differ between groups: CPSP median (min-max) = 4 (1-31), NPSS median (min-max) = 4 (1-8) days, *p* = 0.3587 (W = 386). Although statistically significant differences were observed between the groups on the NIHSS *p*= 0.004, mRS *p*= 0.001, and Barthel Index *p*= <0.001, the absolute scores were low, suggesting the clinical impact of these differences is likely minimal. Further clinical details and statistics for the whole cohort can be found in the supplementary material Table S1.

**Table 1:** Demographic information.

| Dependent: Diagnosis |  | CPSP | NPSS | Total | p |
| --- | --- | --- | --- | --- | --- |
| Total N (%) |  | 20 (30.8) | 45 (69.2) | 65 |  |
| Age (years) | Median (IQR) | 65.0 (58.5 to 68.5) | 64.0 (54.0 to 69.0) | 65.0 (57.0 to 69.0) | 0.787 |
| Sex | Female | 11 (55.0) | 13 (28.9) | 24 (36.9) | 0.055 |
|  | Male | 9 (45.0) | 32 (71.1) | 41 (63.1) |  |
| Thrombolysis | No | 13 (65.0) | 37 (82.2) | 50 (76.9) | 0.201 |
|  | Yes | 7 (35.0) | 8 (17.8) | 15 (23.1) |  |
| Aetiology | Haemorrhagic | 1 (5.0) | 2 (4.4) | 3 (4.6) | 1.000 |
|  | Ischemic | 19 (95.0) | 43 (95.6) | 62 (95.4) |  |
| Lesion side | Bilateral | 1 (5.0) | 1 (2.2) | 2 (3.1) | 0.629 |
|  | Left | 8 (40.0) | 22 (48.9) | 30 (46.2) |  |
|  | Right | 11 (55.0) | 22 (48.9) | 33 (50.8) |  |
| Lesion location | Brainstem | 5 (25.0) | 4 (8.9) | 9 (13.8) | 0.368 |
|  | Cortex | 3 (15.0) | 8 (17.8) | 11 (16.9) |  |
|  | Thalamus | 12 (60.0) | 32 (71.1) | 44 (67.7) |  |
|  | Pathways |  | 1 (2.2) | 1 (1.5) |  |
| Acute volume(ml) | Median (IQR) | 0.6 (0.1 to 1.2) | 0.3 (0.1 to 0.7) | 0.3 (0.1 to 0.8) | 0.343 |
| First available NIHSS | Median (IQR) | 3.0 (1.8 to 5.0) | 2.0 (1.0 to 2.0) | 2.0 (1.0 to 3.0) | 0.004 |
| First available mRS | Median (IQR) | 2.5 (1.0 to 3.2) | 1.0 (1.0 to 2.0) | 1.0 (1.0 to 2.0) | 0.001 |
| First available Barthel index | Median (IQR) | 90.0 (60.0 to 100.0) | 100.0 (100.0 to 100.0) | 100.0 (90.0 to 100.0) | <0.001 |

CPSP, Central post-stroke pain; NPSS, Non-pain sensory stroke; NIHSS, national institutes of health stroke scale; mRS, modified Rankin scale; IQR, interquartile range.

In the acute setting before pain development patients who went on to develop CPSP showed lower grey matter concentration in the contralesional caudate nucleus and nucleus accumbens (NAc) (*t* = 4.17, *p*_FWE-corr_ = 0.021) and in the ipsilesional caudate and NAc (*t* = 4.55, pFWE_-corr_ = 0.020) compared to NPSS patients (Figure 1a: VBM). In the chronic setting (“with pain”) the contralesional caudate nucleus and NAc (*t* = 3.85, *p*_FWE-corr_ = 0.005) showed lower grey matter concentration in CPSP patients (Figure 1b: VBM).

**Figure 1a.**
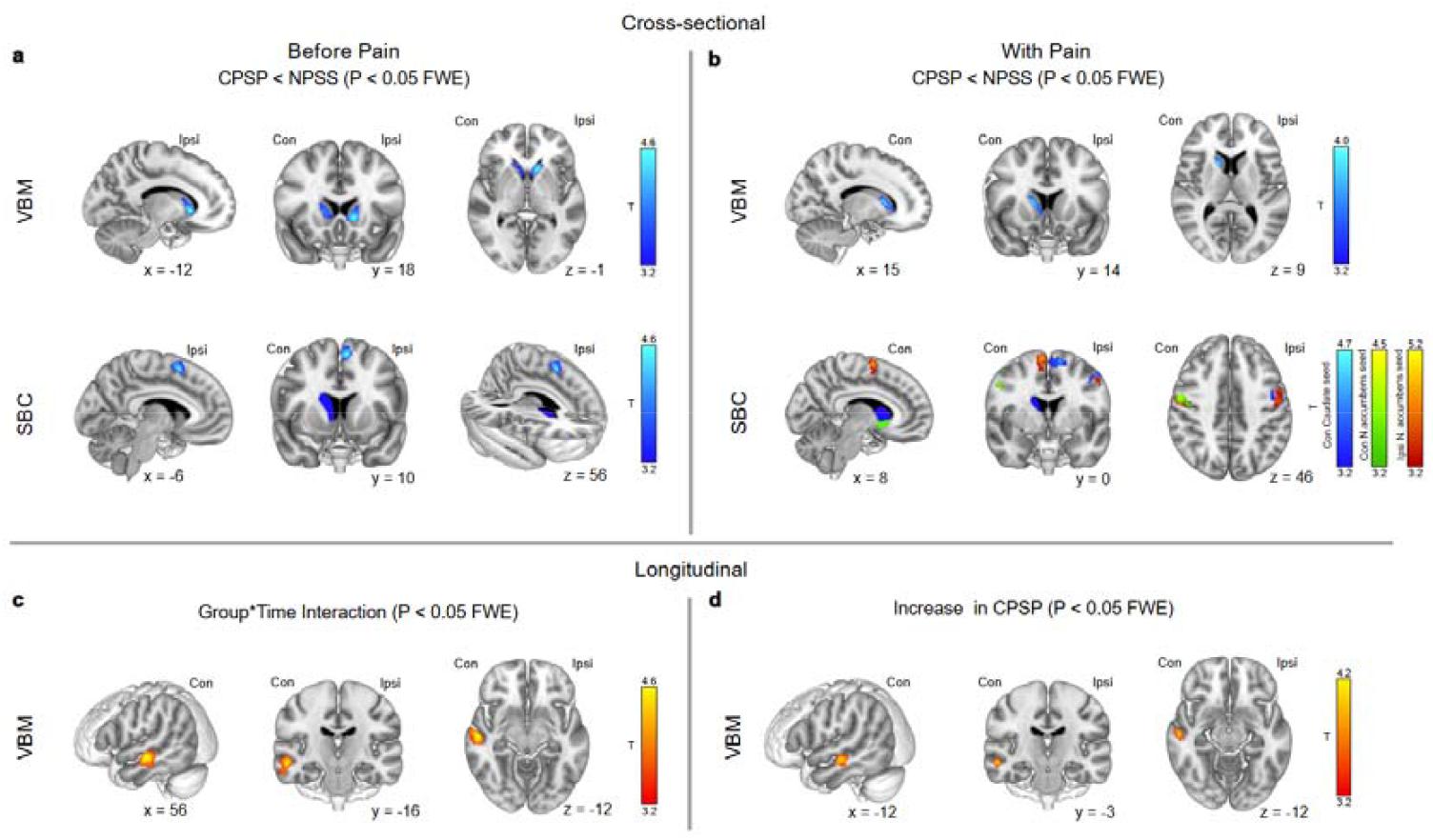
Voxel based morphometry (VBM) analysis of CPSP (central post stroke pain) patients before they develop pain compared to NPSS (non-pain sensory stroke) patients. Cross-sectional analysis in the acute setting with T1 images acquired 2–10 days post stroke. Significantly lower grey matter concentration is seen in stroke patients that go on to develop CPSP when compared to stroke patients that did not develop CPSP in bilateral caudate and NAc. 1a: Seed-based correlation (SBC) analysis in the acute setting before CPSP patients developed pain showing decreased connectivity between the contralesional caudate (seed in blue) and the ipsilesional SMA. 1b: Chronic VBM analysis showing lower grey matter density in the contralesional caudate of CPSP patients after they develop pain compared to NPSS patients.1b: Chronic SBC analysis in CPSP patients the contralesional caudate (blue seed) has decreased connectivity to the ipsilesional SMA. The ipsilesional NAc (red seed) has decreased connectivity to bilateral pre and postcentral gyri and SMAs. The contralesional NAc (green seed) has reduced connectivity to contralesional pre- and postcentral gyri. 1c: Longitudinal VBM analysis showing a significant group and time interaction in the contralesional middle temporal gyrus. 1d: Post-hoc test showing an increase in grey matter density over time in the contralesional middle temporal gyrus only in the CPSP group. T-maps for group comparisons are thresholded at *p* < 0.05 using familywise error correction (FWE) at the cluster level, the extent threshold was chosen to filter out any clusters which did not survive FWE corrections. Sagittal and coronal views of significant clusters with peak voxel coordinates displayed underneath. x, y, z – coordinates in mm; Ipsi – ipsilesional; Con – contralesional.

When comparing changes from the acute phase to the chronic phase, across CPSP and NPSS groups we found a significant interaction between group (CPSP/NPSS) and time (acute/chronic) in the contralesional middle temporal gyrus (*t* = 4.59, *p*_FWE-corr_ = 0.0004) (Figure 1c and Supplementary Table S2). In the CPSP group we see a grey matter density increase over time in the contralesional middle temporal gyrus (*t* = 4.14, *p*_FWE-corr_ = 0.037) (Figure 1d and Supplementary Table S2). In the whole brain analysis, the results were also significant at p<0.001 (see Supplementary material).

In the functional connectivity analysis, we used the regions which showed group differences in the VBM analysis, i.e., the ipsi- and contralesional caudate and NAc as seeds. In the acute setting, the contralesional caudate had a decreased seed-based correlation to the ipsilesional SMA in patients who went on to develop CPSP (*t* = 4.60, *p*_FWE-corr_ = 0.019) (Table 2 and Figure 1a: SBC). The decreased connectivity between the contralesional caudate and the ipsilesional SMA also including the ipsilesional superior frontal gyrus in CPSP patients was confirmed in the chronic phase (with pain) (*t* = 4.55, *p*_FWE-corr_ = 0.018), furthermore, we noted decreased connectivity to the ipsilesional precentral gyrus (*t* = 4.63, *p*_FWE-corr_ = 0.020) (Table 2). For the NAc differences in functional connectivity between CPSP and NPSS patients were noted in the chronic phase only: For the ipsilesional NAc, in CPSP patients, we found reduced functional connectivity to bilateral precentral and postcentral gyri and SMAs and likewise, for the contralesional NAc we noted reduced functional connectivity to the contralesional pre and post central gyri (Table 2 and Figure 1b: SBC).

**Table 2:** MNI coordinates of areas with decreased grey matter concentration and functional connectivity.

|  | **Structural differences CPSP < NPSS acute** |  |  |  |  |  |
| --- | --- | --- | --- | --- | --- | --- |
| **Brain area** | **Cluster level** |  |  | **Peak level** |  | **MNI coordinates** |
| | $P_{\text{FWE corr}}$ | $k_E$ | $P_{\text{uncorr}}$ | $(Z_E)$ | $T$ | mm mm mm |
| Ipsi Caudate | 0.020 | 574 | 0.002 | 4.20 | 4.55 | -12 18 -2 |
| Con Caudate | 0.021 | 564 | 0.002 | 3.89 | 4.17 | 14 12 9 |
|  | **Structural differences CPSP < NPSS chronic** |  |  |  |  |  |
| Ipsi Caudate* | 0.985 | 40 | 0.334 | 3.25 | 3.40 | -14 20 0 |
| Con Caudate | 0.029 | 551 | 0.002 | 3.66 | 3.87 | 15 14 9 |
|  | **Functional differences CPSP < NPSS acute** |  |  |  |  |  |
| Con Caudate - Ipsi SMA | 0.019 | 203 | 0.001 | 4.24 | 4.60 | -6 10 56 |
|  | **Functional differences CPSP < NPSS chronic** |  |  |  |  |  |
| Ipsi accumbens -Ipsi precentral G | 0.002 | 301 | 0.000 | 4.67 | 5.11 | -34 -14 36 |
| Ipsi accumbens- Con precentral G | 0.000 | 465 | 0.000 | 4.53 | 4.92 | 50 -6 50 |
| Ipsi accumbens- Con SMA | 0.008 | 237 | 0.000 | 4.16 | 4.47 | 8 0 66 |
| Con Caudate - Ipsi SMA | 0.018 | 202 | 0.001 | 4.23 | 4.55 | -6 4 56 |
| Con Caudate - Ipsi precentral G | 0.020 | 196 | 0.001 | 4.29 | 4.63 | -46 -2 46 |
| Con accumbens – Con precentral G | 0.003 | 298 | 0.000 | 4.21 | 4.53 | 46 -4 54 |

CPSP central post stroke pain; NPSS non-pain sensory stroke; FWE family wise error correction; SMA supplementary motor area; MNI Montreal Neurological Institute. All right-sided lesions were
flipped to the left side. R caudate (contralesional caudate) was used as the seed. \*The ipsilesional caudate is only significant at the $p < 0.01$ uncorrected level in the chronic phase.

The group-by-time interaction analysis revealed no significant changes over time in the functional connectivity of the caudate nuclei. Using the ipsi- and contralesional NAc as seeds the group-by-time interaction analysis revealed significant group differences in connectivity to the ipsilesional precentral and ipsilesional superior temporal gyrus over time, however, this could not be attributed to a specific group as the post-hoc tests did not identify significant pairwise differences.

The main new findings of our study are structural and functional connectivity changes in CPSP patients already before pain onset, specifically, grey matter decreases in bilateral caudate and NAc and a corresponding decrease in functional connectivity between the contralesional caudate and the ipsilesional SMA. In the chronic phase, structural changes in contralesional caudate and NAc persisted, alongside a decrease in functional connectivity between the bilateral NAc and the sensorimotor cortices.

The *caudate nuclei* are involved in pain modulation and suppression of painful stimuli [9]. Animal models have shown that dopamine agonists and antagonists injected into the rat striatum alter the animal’s nociceptive behaviour [10] and in a PET study in healthy humans, dopamine D2 receptor binding potential in the striatum was shown to play an important role in modulating individual responses to cold pain and central pain regulation [11]. Also, higher availability of dopamine D2/D3 receptors is associated with an enhanced ability to activate pain inhibitory mechanisms that mitigate pain at a remote site [12]. Functional MRI studies have shown that the caudate nuclei are activated during tasks that require healthy participants to judge the location of the painful stimulus and during the anticipation of a painful stimulus [13, 14]. The NAc has been shown to provide negative reinforcement during pain relief [15], and it is part of a network of brain regions that explain trial-by-trial variations in pain ratings beyond fMRI markers of nociception and the pain-modulating effects of expectancy and perceived control [16]. It has also been shown to be involved in placebo analgesia [17].

In addition to these findings, structural and functional changes in the caudate nuclei and/or NAc have been shown in patients with chronic pain [18, 19], specifically in patients with CPSP [7] and with multiple sclerosis-associated central neuropathic pain [20]. Interestingly, a recent VBM study on an experimental model of CPSP in non-human primates also found a decrease in grey matter in the ipsilesional caudate (in addition to changes in the SII, insula and putamen) [21]. Furthermore, the caudate nuclei have been implicated in other central neuropathic pain syndromes, such as post-stroke complex regional pain syndrome (CRPS), with significant associations observed between post-stroke CRPS and lesions involving the head of the caudate nucleus, the putamen, and white matter tracts in the corona radiata [22].

Crucially, in our study, neural alterations in caudate and NAc were already noted before pain developed. In this context, it is noteworthy that several previous studies point to a role of the reward circuitry in the *transition* to chronic pain. A longitudinal study in rats developing peripheral neuropathic pain showed reduced functional connectivity of the caudate and putamen in the early phase of neuropathic pain development [23]. In humans, functional connectivity between the medial prefrontal cortex and the NAc has been shown to predict pain chronification [18], presumably through an alteration in reward learning mechanisms [24]. It is important to note that these reports refer to peripheral neuropathic pain in animals or to chronic back pain in humans. Mechanisms that lead to chronic pain due to peripheral disease may not translate directly to centrally generated neuropathic pain. Nevertheless, it seems very plausible to hypothesise that our neural findings in the pre-pain phase indicate involvement of the reward circuitry in the subsequent development of chronic pain. This would represent the missing pathophysiological link to symptomatic CPSP, beyond the initial lesion to the spinothalamic tract, which is considered a necessary but not sufficient condition for the development of CPSP [25].

It is important to mention that the acute grey matter concentration (and connectivity) differences between CPSP and NPSS patients observed within the first few days following a stroke may either have pre-existed before the stroke or may be a consequence of the stroke, leading to very fast changes. Indeed, rapid brain plasticity (as assessed by VBM) has been reported to occur within an hour in the striatum after administration of haloperidol, associated with a functional decoupling from the motor cortex [26] or in the primary motor cortex after one hour of brain computer interface training [27]. While pre-existing differences that predispose patients to developing pain before the development of stroke cannot be completely ruled out, it is noteworthy that in the recent VBM study in a model of CPSP in non-human primates a decrease in (ipsilesional) caudate grey matter developed after induction of the stroke [21].

Further supporting their role in CPSP generation is the observation that the caudate and nucleus accumbens exhibit not only structural but also functional alterations. We observed decreased functional connectivity between the contralesional caudate and the ipsilesional SMA before the onset of pain. A role of the premotor and motor cortex in pain has been concluded by many other studies. For example, in CPSP patients with pontine lesions, studies employing PET imaging have shown that the reduced metabolism of the ipsilesional SMA correlated with increased pain intensity [28]. Furthermore, motor cortex stimulation has been well established to reduce pain intensity in patients with neuropathic pain and specifically with CPSP [29], which is suggested to be related to a recently identified connection between M1 and NAc [30]. Lesion network mapping has also implicated the primary motor cortex in CPSP, with studies postulating that disruption of the somato-cognitive action network and nociceptive pathways are key underlying mechanisms [31].

We also observed differences overtime in the middle temporal gyrus specific to our CPSP cohort when compared to patients that did not develop pain during follow-up. This area is of interest as it has already been reported to show a decrease in grey matter volume compared to healthy controls in patients with chronic CPSP [7]. Grey matter in the middle temporal gyrus which is part of the temporo-parietal junction (TPJ) has been shown to be inversely related to pain-related thought suppression scores in patients with chronic back pain [32]. The TPJ may play an important role in directing attention away from pain and keeping undesirable thoughts out of awareness. Abnormalities in this area could thus indicate a dysfunction in cognitive control and increased unwanted thoughts about pain. Mechanistically, it has been suggested that the MTG may impact the regulation of pain inhibition through its functional connection with the periaqueductal grey [32, 33], a primary producer of analgesia. The importance of attention and context in pain modulation has been shown by Ploner et al. [34]. Their study showed increased brain connectivity between the anterior insula and emotional networks in frontoparietal and medial temporal lobe areas when attention and emotions to pain are modulated. Future studies might address a possible pathogenetic role of MTG/TPJ in the development of CPSP for example by assessing the effect of interventions with transcranial stimulation.

The main limitation of this study is the relatively small sample size. The results need to be confirmed in a larger, ideally multicentre, study. Furthermore, the patients included had mostly mild stroke-induced impairment, limiting the generalisability to all stroke patients.

In conclusion, our study demonstrates distinct structural and functional differences between CPSP and NPSS patients before pain and during pain development. Alterations in caudate / accumbens preceding pain onset suggest a role of the reward circuitry in the generation of CPSP. Potential preventive interventions might target this circuitry e.g. by pharmacological modulation of dopaminergic neurotransmission, or by transcranial stimulation of the motor system (M1 or SMA) which – via the “M1–accumbens-pathway” [30] – might also modulate activity of NAc.

## Online Methods

### Participants

Patients were prospectively recruited at the Department of Neurology and the Centre for Stroke Research, Charité – Universitätsmedizin Berlin, Germany, for this longitudinal cohort study, which was conducted as part of a larger prospective imaging project [35]. Patients admitted with an acute ischemic or haemorrhagic stroke were recruited from the Stroke Unit of the Charité at Campus Benjamin Franklin, Campus Virchow-Klinikum, and/or Charité Campus Mitte between January 2010 and April 2016.

Ethical approval was granted by the Charité university ethics committee (EA4/003/10) on 04.02.2010 and informed consent was obtained from all the patients prior to inclusion. The study was conducted in accordance with the Declaration of Helsinki. Patient data were pseudo-anonymised and stored electronically. The study pre-registration (regarding data analysis) was uploaded on the open science framework (OSF) platform before data analysis: https://osf.io/4jk9t/?view_only=5342fdfac0a84fac93a9d4a873ac65b0.

### Inclusion and exclusion criteria

The main inclusion criteria included: Patient age: 18-85 years, Ischemic stroke in the sensorimotor system, Somatosensory deficit with proven acute stroke, Informed consent.

The main exclusion criteria included: 1. Contraindications for MRI (e.g., pacemaker or metallic implants), 2. Lack of informed consent or withdrawn consent, 3. An infarct area > 50% of the MCA, 4. Multiple acute infarcts in multiple brain areas, 5. Older infarcts in sensorimotor relevant brain regions, 6. Multiple infarct syndrome and/or marked atrophy or leukoencephalopathy, 7. Severe hemiparesis or hemiplegia, 8. Hydrocephalus, pre-existing neurodegenerative disease, dementia, 9. Immobile patients, 10. Malignant primary disease, 11. Other severe concomitant diseases. In total, 115 patients with stroke were screened for inclusion, 21 of which were excluded as they did not meet the inclusion criteria (Supplementary Figure 1). Seventeen patients declined further participation and two were lost to follow up (total n = 19).

Included patients were prospectively examined for the development of central neuropathic pain. The final number of patients included in the “per protocol” analysis of “CPSP” vs. “NPSS” was n = 75. Of the 75 participants 26 patients developed central neuropathic pain after stroke and were assigned to the “CPSP” group, whereas 49 patients did not develop pain by the last follow up and were assigned to the “NPSS” group. CPSP was defined according to the criteria of Klit *et al*. as pain arising directly from a lesion of the central somatosensory system confirmed by MRI, affecting the body region corresponding to the somatosensory deficit (e.g., hypoesthesia, paraesthesia). Detailed information on patient assessment procedures is provided in Asseyer & Panagoulas *et al*. In the acute cohort, five patients developed pain prior to the first available MRI and were therefore excluded, and one additional patient had missing data, resulting in a total of 20 CPSP patients included in the acute VBM analysis. In the chronic cohort, all CPSP patients were included except for one individual with missing data, yielding a total of 25 CPSP patients; 19 CPSP patients were included in the longitudinal analysis.

In the NPSS group, three patients had missing data in the acute setting and one was excluded due to extensive white matter hyperintensities (WMH), leaving 45 NPSS patients for the acute analyses. In the chronic setting, one additional patient had missing data and one was excluded for WMH, resulting in 47 NPSS patients included in the chronic and 44 in the longitudinal analyses. In the functional MRI analysis, 64 patients were included in the cross-sectional analysis “before pain” including 19 CPSP and 45 NPSS patients, respectively. A total of 70 patients were included in the cross-sectional analysis “with pain” including 25 CPSP and 45 NPSS. In the longitudinal fMRI analysis in total 60 patients were included 18 CPSP and 42 NPSS.

MRI scans were selected according to standardized temporal criteria: (a) for the acute/subacute phase, the first available MRI acquired before the onset of pain (for CPSP) or the first available MRI after stroke (for NPSS); and (b) for the chronic phase, the last available MRI obtained after the onset of pain (for CPSP) or after stroke (for NPSS). Efforts were made to minimize intergroup variability by matching the MRI acquisition times relative to stroke onset as closely as possible.

Five patients developed pain in the first days following the stroke before the first MRI (these patients could therefore not be included in the “cross-sectional, before pain” analysis). Eight patients developed pain within 30 days, seven patients developed pain between 30-90 days after stroke and five patients developed pain between 90-180 days following stroke and one patient developed pain 240 days after a stroke.

### Study design

#### MRI data acquisition

All images were acquired on a 3-Tesla magnetic resonance tomograph (Magnetom TIM TRIO, Siemens AG, Erlangen, Germany, software versions syngo MR B15, B17 and B19) of the Centre for Stroke Research Charité at Campus Benjamin Franklin. A 20-channel head coil was used throughout the study. High resolution three-dimensional structural T1-weighted magnetization prepared rapid gradient echo (MPRAGE) images (echo time [TE]:2.5 ms, repetition time [TR]: 1900 ms, inversion time [TI]: 900 ms, flip angle: 9°, field of view [FOV]: 56 × 256 mm^2^, matrix size = 256 × 256, 192 slices, slice thickness = 1 mm), were acquired for VBM analysis. T2-weighted FLAIR (Fluid attenuated inversion recovery) (slice thickness: 5 mm, TE: 100 ms, TR: 800 ms, flip angle: 130°, FOV: 220 mm, TI: 2370.5 ms), sequences were performed for structural imaging and classification of the lesions in the somatosensory system. Furthermore, lesions were also confirmed on diffusion weighted imaging (DWI) sequences. Additional imaging sequences included diffusion tensor imaging (DTI) and resting state BOLD (rBOLD) imaging (slice thickness: 4 mm, TE: 30 ms, TR: 2300 ms, flip angle: 90°, matrix: 64 × 64, voxel size: 3 × 3 × 4 mm^3^).

#### Clinical data and assessment

For a detailed reporting of the clinical assessment and classification of patients we refer to Asseyer, Panagoulas et al. 2025 [8]. The examination included medical history (including age, sex, risk factors of stroke, and sensory symptoms), clinical neurological examination and quantitative sensory testing (QST) [36, 37]. These examinations were carried out at five timepoints: (i) day one to three, (ii) day four to ten, (iii) one month, (iv) three months, and (v) six months after onset of the stroke.

#### Preprocessing of MRI data

All MRI data were converted from DICOM (Digital Imaging and Communications in Medicine) to NIFTI (Neuroimaging Informatics Technology Initiative) images using mricron (v1.0.20190902) [38]. ITK-SNAP (3.6.0) was used by an experienced neuroradiologist (KV) to delineate all lesions[39]. Lesion volumes were manually traced on acute MRI scans (DWI/FLAIR) as well as on day 30 (T1 and FLAIR) scans. For 12 subjects, later scans at day 60 or 90 were used when the 30-day images were not available. Structural images of patients with lesions on the right side were flipped from right to left using FSL (5.0.11) to make sure all lesions were located on the same (left) side.

The SPM12 (Statistical Parametric Mapping, London, UK) and CAT12 (Computational Anatomy) toolboxes were used for the VBM analysis and the Functional Connectivity Toolbox (CONN 20b) for the functional connectivity analysis [40]. The T1 images were pre-processed using the longitudinal segmentation pipeline of the CAT12.8.1 (2043) toolbox. The longitudinal model optimized for detecting small changes (i.e. plasticity / learning effects) was used. During the longitudinal pipeline the MRI scans from the different time points of each subject are realigned together and bias-field corrected before they are processed individually. The T1 MPRAGE flipped and lesion masked images were denoised, bias field corrected, segmented, and normalised to MNI space. Furthermore, the grey matter and white matter affine transformed tissue segments were exported to be used with DARTEL (Diffeomorphic Anatomical Registration Through Exponentiated Lie Algebra) to create a customised template from our study sample, which was used during normalisation [41]. Unmodulated images were used in our analysis, as previous studies have demonstrated that, when combined with high-accuracy alignment algorithms, they are sensitive to mesoscopic structural changes [42].

All images were smoothed using a Gaussian kernel of 8 mm^3^ full width at half-maximum (FWHM). Cost function masking was used during the normalisation procedures to reduce deformation due to the stroke lesions [43]. Quality control was performed by looking at the CAT12 quality report parameters, checking the sample homogeneity as well as visually inspecting the pre-processed images. Scans with a quality rating of D and below were rejected.

The resting-state functional data underwent standard preprocessing, which included: functional realignment and unwarping, slice-timing correction, outlier detection via ART-based scrubbing, and segmentation of structural and functional images into grey matter (GM), white matter (WM), and cerebrospinal fluid (CSF) tissue maps. The data were then normalised to the Montreal Neurological Institute (MNI) space, and functional images were smoothed using a Gaussian kernel of 8-mm FWHM. Denoising was performed using CONN’s default settings. Confounding effects, including WM (five regressors), CSF (five regressors), realignment parameters (24 regressors), scrubbing results (22 regressors), and the resting-state measurement (two regressors), were regressed out of the fMRI time series. Finally, the functional data were band-pass filtered (0.008-0.2 Hz) and linearly detrended.

### Statistical analysis

#### Clinical Data

The Fisher exact test was used for categorical variables, and the non-parametric Mann-Whitney U test was used for all other variables. The software package “R studio” (R version 4.2.2) was used for the statistical analysis of demographic and behavioural data.

#### VBM

Statistical analysis was conducted using SPM12. The smoothed segmented and normalised grey matter images from the first and last available visits were included for each patient. A cross-sectional baseline comparison was performed between patients who later developed pain (CPSP) but were still pain-free at baseline (when the first MRI was performed) and those who remained pain-free during follow-up (NPSS). Grey matter maps were compared between NPSS and CPSP groups in both the acute and chronic stages using a two-sample *t*-test, with age, sex, and total intracranial volume (TIV) included as nuisance covariates.

T-maps were log-transformed and labelled using the AAL3 atlases, available from the CAT12 toolbox. A grey matter mask with an absolute threshold of 0.1 was applied to the data to ensure that the analysis is confined to grey matter segments [40]. Exploratory whole brain analyses were performed with an uncorrected threshold of *p* <.001 [7]. Statistically significant results were corrected for multiple comparisons with FWE and an alpha level of 0.05 was chosen. Furthermore, to map structural changes over time a longitudinal analysis was conducted using the flexible factorial design with the CAT12 toolbox.

In our statistical design we specified three factors: subject, group (CPSP, NPSS), and time (pre, post). Here we tested for a potential interaction between group and time to investigate whether there is a significant group difference (CPSP vs. NPSS) related to a grey matter concentration change over time.

Post-hoc paired t-tests were run to investigate whether one group drove the interaction effect. Additionally, a region of interest analysis was conducted using the regions of GMC decrease as reported by Krause and colleagues [7] (See supplementary material).

### Resting state functional connectivity

Seed-to-voxel connectivity analyses were conducted using the left and right caudate and NAc as seed regions as defined in [44]. Further whole brain seed-based correlation analysis as well as connectivity gradient analysis were conducted and will be reported by Chen et al. (in preparation).

BOLD time series were extracted for all voxels and first-level correlation maps were generated by calculating Pearson’s correlation coefficients between the seed time series and those of the other voxels. These correlation coefficients were then transformed into Z-scores using Fisher’s transformation. Two-sample t-tests were performed to assess differences in functional connectivity (FC) between the CPSP and NPSS groups, as well as before and after the onset of pain in CPSP patients. In addition, we also investigated potential group differences over time using a group and time interaction analysis and follow up post-hoc t-tests. For the second-level analyses, an uncorrected threshold of *p* < 0.001 was used, with significant results corrected for FWE at *p* < 0.05.

## Data Availability

Data and the analysis code are available upon reasonable request.

## Acknowledgements

We are deeply grateful to our colleagues in the Stroke Imaging and Trial Teams at the Centre for Stroke Research Berlin (CSB), as well as those in the Department of Neurology, for their dedicated work in recruiting participants, conducting the study, and gathering the data. The authors note that ChatGPT and DeepL assisted with refining the paper’s grammar and phrasing. We also extend our appreciation to Joshua Grant for his careful proofreading of the final manuscript, and, above all, to the patients whose participation made this study possible.

## Supplementary material

Supplementary material are available online.

## Footnotes

1 In this study we looked at the functional connectivity of the structurally affected regions that were significant in the VBM analysis. Further whole brain seed-based correlation analysis as well as connectivity gradient analysis were conducted and will be reported by Chen et al (in preparation).

