## Supplementary Figure 1 for "Reduced Grey Matter in Caudate and Accumbens Nuclei Precedes Central Post Stroke Pain"

Clinical features and demographics

In total 75 patients recruited at the Department of Neurology and the Centre for Stroke Research; Charité-Universitätsmedizin Berlin were included in the analysis and prospectively followed up for the development of pain. Twenty-six of the 75 patients developed CPSP and 49 did not develop CPSP during the follow up period and so were assigned to the NPSS group. CPSP and NPSS patients did not differ significantly in terms of age however there was a significant difference in terms of sex *P*=.028. In the NPSS group 30.6% of the patients were female, whereas in the CPSP 57.7% of patients were female. No significant differences were found for thrombolysis, stroke risk factors (apart from hypercholesterolemia), lesion location, and acute lesion volume between CPSP and NPSS patients. The two patient groups were compared in the acute phase in terms of stroke severity using the NIHSS score, Barthel index and mRS, and in terms of quality of life using the PSQI, SF12 and GDS scales. In the acute phase CPSP patients were more neurologically impaired as indicated by the NIHSS (P_NIHSS_=0.001), mRS score (P_mRS_=0.001) and Barthel index (P_Barthel_=0.002) and had a poorer quality of life in the physical component summary of the SF-12 scale (P_SF12_PCS_=0.025) compared to NPSS patients. There was no significant difference between depression and sleeping quality in the acute phase between the two groups. In the chronic phase CPSP patients were more severely affected in NIHSS (P_NIHSS_=0.001) and mRS scores (P_mRS_<0.001), however it is important to note that both groups had very low scores suggesting that they were very mildly affected, therefore we do not think this substantially contributes to group differences. There was no significant difference between Barthel index and GDS depression scores. CPSP patients did score lower in terms of sleep quality (P_PSQI_=0.019) and in terms of the physical component summary (P_SF12_PCS_=0.021) (Supplementary Table S1).

Supplementary Table S1

| Dependent: Diagnosis |  | CPSP | NPSS | Total | p |
| --- | --- | --- | --- | --- | --- |
| Total N (%) |  | 26 (34.7) | 49 (65.3) | 75 |  |
| Age (years) | Median (IQR) | 63.0 (55.5 to 69.5) | 65.0 (56.0 to 70.0) | 65.0 (55.5 to 70.0) | 0.660 |
| Sex | Female | 15 (57.7) | 15 (30.6) | 30 (40.0) | 0.028 |
|  | Male | 11 (42.3) | 34 (69.4) | 45 (60.0) |  |
| Thrombolysis | No | 18 (69.2) | 40 (81.6) | 58 (77.3) | 0.255 |
|  | Yes | 8 (30.8) | 9 (18.4) | 17 (22.7) |  |
| Aetiology | Haemorrhagic | 1 (3.8) | 2 (4.1) | 3 (4.0) | 1.000 |
|  | Ischemic | 25 (96.2) | 47 (95.9) | 72 (96.0) |  |
| Lesion side | Bilateral | 1 (3.8) | 1 (2.0) | 2 (2.7) | 0.406 |
|  | Left | 9 (34.6) | 24 (49.0) | 33 (44.0) |  |
|  | Right | 16 (61.5) | 24 (49.0) | 40 (53.3) |  |
| Lesion location | Brainstem | 6 (23.1) | 5 (10.2) | 11 (14.7) | 0.191 |
|  | Cortex | 7 (26.9) | 8 (16.3) | 15 (20.0) |  |
|  | Thalamus | 13 (50.0) | 35 (71.4) | 48 (64.0) |  |
|  | Pathways |  | 1 (2.0) | 1 (1.3) |  |
| Acute volume(ml) | Median (IQR) | 0.6 (0.2 to 1.3) | 0.3 (0.1 to 0.5) | 0.3 (0.1 to 0.9) | 0.065 |
| Hypertension | No | 8 (30.8) | 8 (16.3) | 16 (21.3) | 0.235 |
|  | Yes | 18 (69.2) | 41 (83.7) | 59 (78.7) |  |
| Diabetes | No | 23 (88.5) | 42 (85.7) | 65 (86.7) | 1.000 |
|  | Yes | 3 (11.5) | 7 (14.3) | 10 (13.3) |  |
| Smoking | No | 19 (73.1) | 35 (71.4) | 54 (72.0) | 1.000 |
|  | Yes | 7 (26.9) | 14 (28.6) | 21 (28.0) |  |
| Hypercholesterolemia | No | 2 (7.7) | 16 (32.7) | 18 (24.0) | 0.022 |
|  | Yes | 24 (92.3) | 33 (67.3) | 57 (76.0) |  |
| AF | No | 22 (84.6) | 43 (87.8) | 65 (86.7) | 0.731 |
|  | Yes | 4 (15.4) | 6 (12.2) | 10 (13.3) |  |
| BMI > 30 | No | 22 (84.6) | 46 (93.9) | 68 (90.7) | 0.227 |
|  | Yes | 4 (15.4) | 3 (6.1) | 7 (9.3) |  |
| Family history | No | 25 (96.2) | 42 (85.7) | 67 (89.3) | 0.249 |
|  | Yes | 1 (3.8) | 7 (14.3) | 8 (10.7) |  |
| First available NIHSS | Median (IQR) | 3.0 (2.0 to 5.0) | 2.0 (1.0 to 2.0) | 2.0 (1.0 to 3.0) | 0.001 |
| First available mRS | Median (IQR) | 2.0 (1.0 to 3.8) | 1.0 (1.0 to 2.0) | 1.0 (1.0 to 2.0) | 0.001 |
| First available Barthel index | Median (IQR) | 90.0 (61.2 to 100.0) | 100.0 (100.0 to 100.0) | 100.0 (90.0 to 100.0) | 0.002 |
| Acute PSQI | Median (IQR) | 6.0 (3.0 to 7.0) | 4.0 (2.0 to 6.0) | 5.0 (3.0 to 7.0) | 0.078 |
| Acute SF-12-MCS | Median (IQR) | 55.8 (48.4 to 60.8) | 55.9 (50.7 to 58.4) | 55.9 (50.3 to 59.0) | 0.504 |
| Acute SF-12-PCS | Median (IQR) | 45.0 (42.5 to 52.9) | 51.7 (47.4 to 54.2) | 50.3 (44.6 to 54.0) | 0.025 |
| Acute GDS | Median (IQR) | 5.0 (2.0 to 8.0) | 4.0 (1.0 to 7.0) | 4.0 (1.0 to 8.0) | 0.557 |
| Chronic NIHSS | Median (IQR) | 1.0 (1.0 to 2.0) | 1.0 (0.0 to 1.0) | 1.0 (0.0 to 1.0) | 0.001 |
| Chronic mRS | Median (IQR) | 1.0 (1.0 to 2.0) | 1.0 (1.0 to 1.0) | 1.0 (1.0 to 1.0) | <0.001 |
| Chronic Barthel index | Median (IQR) | 100.0 (100.0 to 100.0) | 100.0 (100.0 to 100.0) | 100.0 (100.0 to 100.0) | 0.239 |
| Chronic PSQI | Median (IQR) | 6.0 (3.2 to 8.8) | 4.0 (3.0 to 5.0) | 4.0 (3.0 to 6.5) | 0.019 |
| Chronic SF-12-MCS | Median (IQR) | 55.9 (53.5 to 58.8) | 55.9 (50.2 to 57.8) | 55.9 (51.4 to 58.2) | 0.581 |
| Chronic SF-12-PCS | Median (IQR) | 50.2 (40.7 to 53.8) | 53.6 (47.2 to 55.5) | 53.1 (45.6 to 54.6) | 0.021 |
| Chronic GDS | Median (IQR) | 4.5 (2.0 to 7.8) | 4.0 (2.0 to 7.0) | 4.0 (2.0 to 7.0) | 0.447 |

Longitudinal and region of interest analysis

The areas with significant loss of grey matter density in both groups are the medial right and left postcentral gyrus. This is in line with our expectations as patients suffered from a somatosensory stroke. The following areas were used as regions of interest from the Krause et al. 2016 paper which found significant grey matter volume decreases in pain patients in comparison to healthy controls in: ipsilesional and contralesional S2, ipsilesional anterior and posterior insular cortex, ipsilesional ventrolateral prefrontal cortex, contralesional anterior insular cortex, contralesional nucleus Accumbens, contralesional orbitofrontal cortex, contralesional medial temporal gyrus and superior temporal gyrus. Interestingly we see increases over time in the contralesional middle temporal gyrus in the CPSP group compared to the NPSS group (p_FEWcorr_=0.037, T-value=4.14).

Supplementary Table S2 ROI analysis Longitudinal

| **Longitudinal results for both groups showing decrease in Grey matter density over the post central gyrus** | | | | | |
| --- | --- | --- | --- | --- | --- |
| P_FWE_ | P_uncorr_ | T value | Cluster size | MNI coordinates | Anatomical area aal3 Atlas |
| 0.000 | 0.000 | 5.91 | 4025 | -3 -45 66 | Ipsi Postcentral gyrus |
|  | |  |  |  | Ipsi Superior parietal lobule |
|  | |  |  |  | Ipsi Precuneus |
| **Time x Group interaction showing grey matter increase over time in the contralesional middle temporal gyrus** | | | | | |
| 0.0004 | 0.00005 | 4.59 | 1155 | 56 -20 -10 | Con Mid Temporal G |
| **Post-hoc test showing grey matter increase in the contralesional middle temporal gyrus specific to the pain group** | | | | | |
| 0.037 | 0.008 | 4.14 | 424 | 56 -20 12 | Con Mid Temporal G |

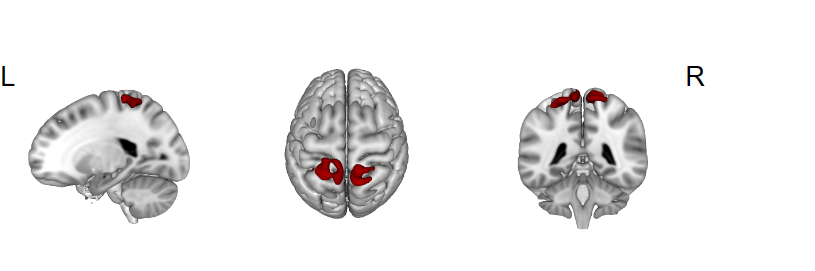

Supplementary Figure S2 Grey matter concentration loss in bilateral SI in patients with somatosensory stroke.
